# SARS-CoV-2 antibody prevalence in health care workers: Preliminary report of a single center study

**DOI:** 10.1101/2020.07.20.20158329

**Authors:** Michael Brant-Zawadzki, Deborah Fridman, Philip A. Robinson, Matthew Zahn, Randy German, Marcus Breit, Junko Hara

## Abstract

SARS-CoV-2 has driven a pandemic crisis. Serological surveys have been conducted to establish prevalence for covid-19 antibody in various cohorts and communities. However, the prevalence among healthcare workers is still being analyzed. The present study reports on initial sero-surveillance conducted on healthcare workers at a regional hospital system in Orange County, California, during May and June, 2020.

Study participants were recruited from the entire hospital employee workforce and the independent medical staff. Data were collected for job title, location, covid-19 symptoms, a PCR test history, travel record since January 2020, and existence of household contacts with covid-19. A blood sample was collected from each subject for serum analysis for IgG antibodies to SARS-CoV-2.

Of 3,013 tested individuals, a total 2,932 were included in the analysis due to some missing data. Observed prevalence of 1.06% (31 antibody positive cases), adjusted prevalence of 1.13% for test sensitivity and specificity were identified. Significant group differences between positive vs. negative were observed for age (z = 2.65, *p* = .008), race (*p* = .037), presence of fever (*p* < .001) and loss of smell (*p* < .001).

Possible explanation for this low prevalence includes a relatively low local geographic community prevalence (∼4.4%) at the time of testing, the hospital’s timely procurement of personal protective equipment, rigorous employee education, patient triage and treatment protocol development and implementation. In addition, possible greater presence of cross-reactive adaptive T cell mediated immunity in healthcare workers vs. the general population may have contributed. Determining antibody prevalence in front-line workers, and duration of antibody presence may help stratify the workforce for risk, establish better health place policies and procedures, and potentially better mitigate transmission.

## Introduction

The SARS-CoV-2 pandemic crisis is evolving. Its hallmark is very high infectivity, pre-symptomatic transmission and asymptomatic prevalence which continue to fuel dramatic cumulative numbers of infections, hospitalizations, and deaths. To better understand the extent of undetected transmission, serological surveys in sampled cohorts identified antibodies from prior infection ranging from 57% prevalence in Bergamo - Italy’s epicenter [1], 20% in New York City [2], down to 4.7 % in Los Angeles County [3] and 2.8% in Santa Clara County [4], California.

Prevalence of antibodies among healthcare workers, presumed at higher risk for infection, has not been well-established. Determining such prevalence in front-line workers, and duration of antibody presence may help stratify the workforce for risk, establish better health place policies and procedures, and potentially better mitigate transmission.

This article reports on initial sero-surveillance conducted in 3,013 healthcare workers at Hoag Memorial Hospital Presbyterian, California, United States, during May and June, 2020. IRB approval was obtained for this study (Providence St. Joseph Health IRB # 2020000337).

## Methods

Study participants were recruited by email notifications to the entire employee workforce (6000+ individuals) and the independent medical staff (1600+ physicians). The consenting participants were interviewed as to job title, location, covid-19 symptoms, a PCR test history, travel record since January 2020, and existence of household contacts with covid-19 outside of work. A blood sample (∼5ml) was collected from each subject for serum analysis for IgG antibodies to SARS-CoV-2 using the VITROS Anti-SARS-CoV-2 IgG Reagent Pack and Calibrator on the VITROS® XT 7600 instrument by Ortho Clinical Diagnostics.

## Results

Of an initial 3,013 samples recorded, subjects were excluded from analyses due to missing age (n = 24), gender (n = 14), race, (n = 31), and symptoms (n = 12), resulting in a complete pool of 2,932 (**Table 1**). Antibody testing identified 31 positive cases (2,901 negative), thus an observed prevalence of 1.06% (exact binomial 95% CI = 0.71% - 1.50%). Accounting for test sensitivity of 93.6% and specificity of 100%, an adjusted prevalence of 1.13% (95% CI = 0.78% - 1.58%) was calculated, indicating 33 positive cases (negative = 2,899) after adjustment.

**Table 1.** Sample Characteristics and Group Differences.

|  | Antibody<br>Negative | Antibody<br>Positive | Total |  |
| --- | --- | --- | --- | --- |
| | n = 2901<br>(99%) | n = 31<br>(1%) | N = 2932<br>(100%) | $p^a$ |
| Age in yrs., *M (SD)* | 42.67 (12.10) | 37.58 (12.30) | 42.62 (12.12) | .008 |
| Female, count (%) | 2102 (72%) | 23 (74%) | 2125 (72%) | .507 |
| Race, count (%) |  |  |  | .037 |
| American Indian or Alaska Native | 19 (1%) | 0 | 19 (1%) |  |
| Asian | 656 (23%) | 10 (32%) | 666 (23%) |  |
| Black | 47 (2%) | 0 | 47 (2%) |  |
| Hispanic or Latino | 486 (17%) | 11 (35%) | 497 (17%) |  |
| Native Hawaiian or Pacific Islander | 50 (2%) | 1 (3%) | 51 (2%) |  |
| White | 1459 (50%) | 9 (29%) | 1468 (50%) |  |
| Other | 184 (6%) | 0 | 184 (6%) |  |
| Fever, count (%) | 331 (11%) | 12 (39%) | 343 (12%) | < .001 |
| Cough, count (%) | 474 (16%) | 7 (23%) | 481 (16%) | .332 |
| Sore Throat, count (%) | 550 (19%) | 7 (23%) | 557 (19%) | .644 |
| Runny Nose, count (%) | 403 (14%) | 7 (23%) | 410 (14%) | .187 |
| Loss of Smell, count (%) | 55 (2%) | 13 (42%) | 68 (2%) | < .001 |

<sup>a</sup> Group difference testing was performed with Mann-Whitney U tests for age and with Fisher's exact tests for categorical measures.

Nonparametric tests for group differences were performed for demographics and five symptoms of covid-19. Significant differences between observed negative and positive cases were found for age (z = 2.65, *p* = .008), race (*p* = .037), presence of fever (*p* < .001), and loss of smell (*p* < .001). Of those with previously confirmed diagnosis of covid-19 (n = 12), 6 were antibody positive with 6 non-reactive.

## Discussion

The initial result found a significantly lower prevalence of SARS-CoV-2 antibody carriers among our healthcare workers compared to prior reports. During this same period, our prevalence of antibodies tested by physician order in our community was 3.87%.

One possible explanation is a relatively low regional estimated prevalence of infections (∼4.4%) further evidenced by average 104 patients per day in ICU and 330 cumulative death in Orange County (total population of 3.18 million) at the time of our study. Also, our institution had implemented stringent workforce education on personal hygiene, social distancing and appropriate PPE usage since January 2020 when we saw the first California and 3rd US case, with hospital-wide protocols in patient triage, and symptom surveillance. Finally, recent research suggests possibly greater presence of cross-reactive adaptive T cell mediated immunity in healthcare workers vs the general population. Several studies have documented such innate T-cell immunity can exist related to prior exposure of similar Corona virus exposure, and cross-reactivity such related virus species. It is assumed workplace exposure is more frequent for health care workers to such various coronavirus pathogens [5,6]. A combination of these factors may explain our findings.

We will retest this same cohort at 8 weeks and 6 months, to better understand the dynamics of SARS-CoV-2 antibody prevalence and duration in healthcare workers.

## Data Availability

Data cannot be shared publicly because of possible hospital data access restrictions. However, upon acceptance of the manuscript, data will be available from the Hoag Center for Research and Education (contact via) for researchers who meet the criteria for access to confidential data.

## Acknowledgements

We acknowledge our healthcare workers who have contributed to this study. We also thank Mr. Jason Bock from Medical Care Corporation for his assistance in data analyses. This study is funded by Orange County Healthcare Agency and Hoag Hospital Foundation.

## Notes

### Competing Interest Statement

The authors have declared no competing interest.

### Author Declarations

IRB approval was obtained for this study from Providence St. Joseph Health IRB (# 2020000337).

